# Correcting under-reported COVID-19 case numbers: estimating the true scale of the pandemic

**DOI:** 10.1101/2020.03.14.20036178

**Authors:** Kathleen M. Jagodnik, Forest Ray, Federico M. Giorgi, Alexander Lachmann

## Abstract

The COVID-19 virus has spread worldwide in a matter of a few months, while healthcare systems struggle to monitor and report current cases. Testing results have struggled with the relative capabilities, testing policies and preparedness of each affected country, making their comparison a non-trivial task. Since severe cases, which more likely lead to fatal outcomes, are detected at a higher rate than mild cases, the reported virus mortality is likely inflated in most countries. Lockdowns and changes in human behavior modulate the underlying growth rate of the virus. Under-sampling of infection cases may lead to the under-estimation of total cases, resulting in systematic mortality estimation biases. For healthcare systems worldwide it is important to know the expected number of cases that will need treatment. In this manuscript, we identify a generalizable growth rate decay reflecting behavioral change. We propose a method to correct the reported COVID-19 cases and death numbers by using a benchmark country (South Korea) with near-optimal testing coverage, with considerations on population demographics. We extrapolate expected deaths and hospitalizations with respect to observations in countries that passed the exponential growth curve. By applying our correction, we predict that the number of cases is highly under-reported in most countries and a significant burden on worldwide hospital capacity.

The full analysis workflow and data is available at: https://github.com/lachmann12/covid19

## Introduction

Coronavirus Disease 19 (COVID-19) is a novel human illness caused by the Severe Acute Respiratory Syndrome CoronaVirus 2 (SARS-CoV-2), a pathogen initially discovered in the Wuhan region of China at the end of 2019 (1). It has reached pandemic status by the World Health Organization (WHO) within less than four months of initial reports of the disease. The origin of the virus can be traced back to related strains predominantly found in bats (2). Individuals infected by the disease can experience a range of symptoms, including cough, chills, fever, and shortness of breath (1). From data currently available, fatal disease progression is higher than that of the common influenza strains, and as such, more COVID-19-related deaths are occurring than had resulted from the recent Severe Acute Respiratory Syndrome (SARS) virus and the Middle East Respiratory Syndrome (MERS) combined (3). The infection rate of COVID-19 has been estimated between an *R*_0_ of 2 and up to 6.49 (4) compared with influenza, characterized by an *R*_0_ of approximately 1.3 (5). The severity of infection is highly correlated to the age of the infected individual. Younger segments of a population present a much lower risk than older populations. A current data release from the Center for Disease Control in South Korea shows that while there are few reported fatalities for individuals under 30 years of age, the case fatality rate (CFR) (percent of deaths per confirmed case) for individuals older than 80 is over 8% (6). Figure 1 shows six countries with a significant number of reported COVID-19 cases. China, which has been the origin of the outbreak (7), initially registered the most cases early on with over 80,000, but has since been overtaken by the US. Through severe containment measures such as curfews and lockdowns of public life, new infections have slowed significantly (8). Other countries that have been affected only recently are still in the exponential growth curve and have at this point overtaken China in the total number of confirmed COVID-19 cases. Countries like Italy and the US have only recently taken action to slow the spread of the virus. With a reported incubation time of about five days, there is a time-delay for the effects of a slowdown to becoe evident (9). Another country that is currently experiencing high numbers of reported COVID-19 cases is Iran, with more than 12,000 confirmed cases. Similar to Italy the growth rates are slowing for more than two weeks as of writing. Due to the limited information available, most parameters describing the dynamics of the disease spread involve significant uncertainties. Healthcare systems in most countries are not capable of monitoring the exponential growth of a virus in this manner. South Korea, as of writing, has one of the most extensive capabilities of testing individuals per capita, with a capacity of more than 20,000 tests a day. Hence, South Korea represents the best benchmark country in order to predict the COVID-19 CFR. The proposed method uses demographic information to identify the fraction of the vulnerable population. Countries such as China have a generally younger population, reducing the overall risk of fatal outcomes, and this should result in a lower CFR compared with South Korea. Countries such as Italy, having an older population compared with South Korea, should have higher CFRs. Estimating the true case count is relevant to identifying the correct measures to stop the disease from spreading and to predict expected hospitalization needs.

**Fig. 1.**
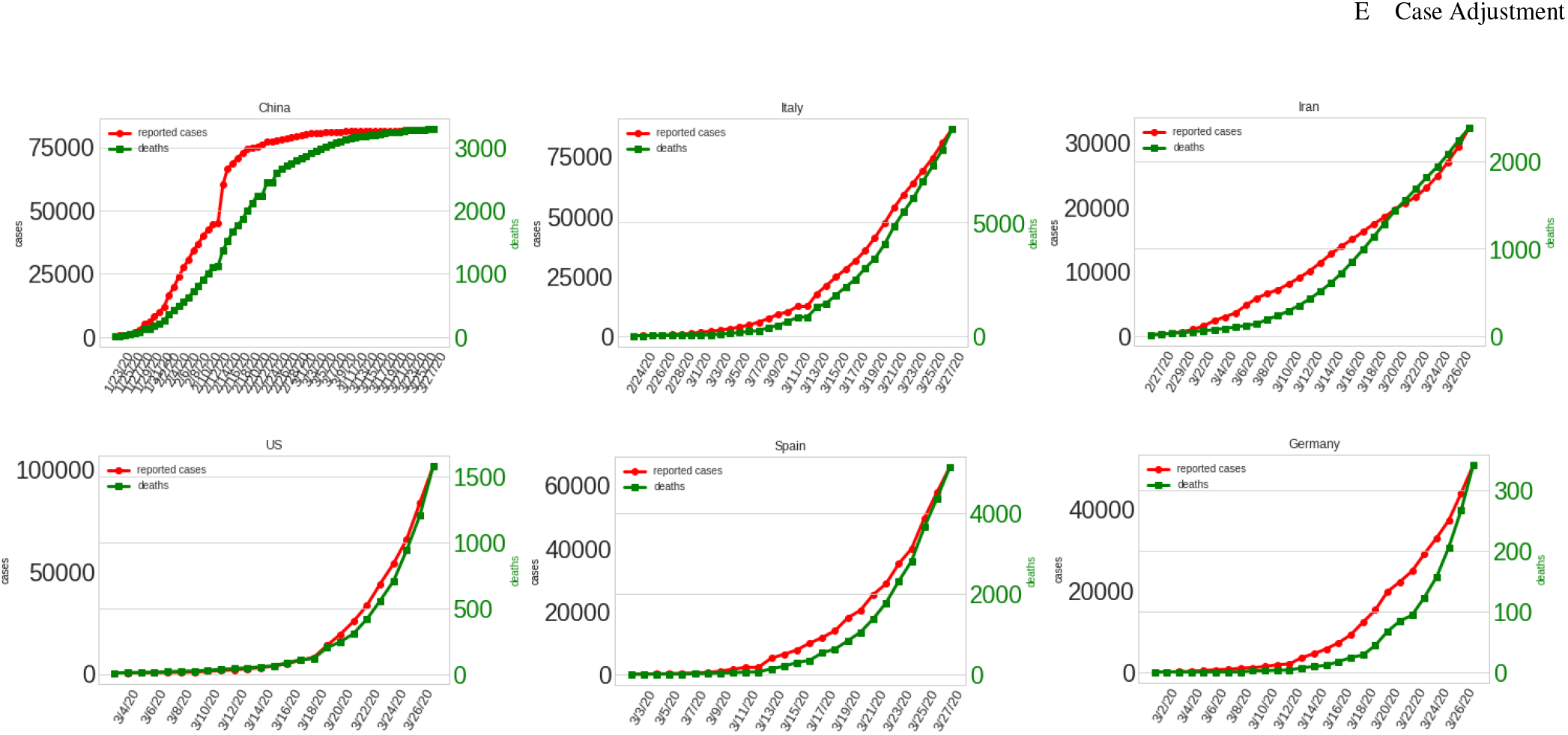
Case progression for 6 countries with high COVID-19 case numbers.

## Methods

### A. Data

The case correction relies on two datasets. The first is the data published by the WHO, which is updated every day and contains case, recovery, and death numbers for countries reporting all known COVID-19 cases (10). The second dataset is a global demographic database maintained by the United Nations (11). This database contains the number of individuals per year of age for more than 200 countries. For the analysis, we extracted the data between 2007 and 2019. We always choose the most recent data entry for the countries if multiple exist. This file is hosted as a Kaggle dataset at:https://www.kaggle.com/lachmann12/world-population-demographics-by-age-2019.

### B. Limitations

This method makes a series of assumptions in order to adjust reported COVID-19 cases compared with the benchmark country (South Korea). As the pandemic is still evolving may parameters are not sufficiently known.

- **Deaths are a proxy measure for COVID-19 cases** We estimate cumulative case numbers purely from reported cumulative deaths. The average time of death after infection os 23 days, so the number of reported deaths at dat *t* are a proxy measure of cases at *t −* 23.
- **Deaths are confirmed equally** It is assumed that if a death occurs due to COVID-19, the case will be confirmed. When there is under-reporting, the reported CFR would be lower than the true CFR.
- **The population is infected uniformly** We assume that the probability of infection is uniformly distributed across all age groups. The probability of an 80-year-old person to become infected is equal to the probability of a 30-year-old to become infected.
- **Changes in healthcare load are not modeled** The provided healthcare in countries is comparable. For developed countries such as Italy and South Korea, it is assumed that the population has similar access to treatment. The CFRs reported by age group are thus applicable in all countries.
- **The virus is identical in all countries** This is supported by the very low mutational rate of SARS-CoV-2, which allows conjecturing identical etiologies across countries (12).
- **Conservative modeling** Our method relies on estimating future cumulative deaths for a period of at least 23 days. In most countries in this study there has been no observed change in growth rate up to the day of writing. The model assumes that the growth rate will start falling on the next day and follow our precomputed spline.
- **The model does not consider population saturation** We propose this method as an early prediction tool to estimate potential hospitalizations and deaths at the early stages of the viral spread. Unlike SIR (4) models the growth rate is not modeled on the percent of the susceptible population.

### C. Calculating growth rate spline modeling behavior change

We use the observed growth rate curves of China, Italy, and Iran to estimate the average growth rate change on population behavior changes and the effect on growth rate over time. We align the growth rate curves for the three countries to achieve a best fit on the exponentially decreasing part of recorded growth rates. The onset of the curves does not have to fall on the days where a lockdown was put in place. We fit the data points from China, Italy, and Iran to a exponentially decreasing growth rate function *EDG*(*t*) with two parameters *a* and *b*.

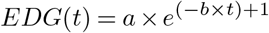

for day *t* after behavior change and constant *a* and exponent factor *b*.To fit t he d ata p oints w e u se the *optimize*.*curve*_*f it* implementation of the *scipy*.*optimize* library (13).

### D. Extrapolating growth rates from reported deaths

The proposed model assumes that the growth of COVID-19 undergoes two distinct phases. The first i s unhindered growth at observed constant rates of 1.33/day. This expansion phase is followed by an exponentially decreasing function. We align countries to the respected position in the growth rate phase. If the latest available growth rates are showing constant growth rates we assume the country will enter the second phase the next day. We estimate predicted deaths at future time points with:

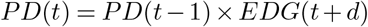

, for *d* representing the alignment of growth rates for the pre-computed growth rate spline and the growth rate development of the country analysed.

### E. Case Adjustment

CFRs have significantly changed over time and country. In some countries the CFR In this context, it can be noted that South Korea and Germany show the most consistent CFR estimates, but are still rising (data not shown). These two countries show a significantly lower CFR compared with other countries. The change of CFR over time within the same country is potentially caused by changes in the number of false-negative cases, meaning that many infections go unnoticed until they become fatal. Additionally, countries undergoing exponential growth have over proportionally new cases. In the case of Italy, there might not have been sufficient capacity to confirm infections. With a smaller fraction of potential cases tested, the estimated CFR will increase. In the case of Italy, the estimated rate increased from 2% to more than 6%.

This method requires the comparison of two countries with sufficient confirmed cases and reported deaths. One country (target country) will be adjusted, given the information from the second country (benchmark country). In order to adjust for the difference in the population demographics of the target country, 𝕋, and the benchmark country, 𝔹, we compute a Vulnerability Factor (*V*_𝕋 𝔹_).

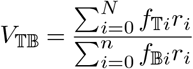

, where *f*_𝕋*i*_ is the fraction of the population with age *i* for target country 𝕋, *f*_B*i*_ is the fraction of the population with age *i* for benchmark country 𝔹, and *r*_*i*_ the CFR for age *i. r*_*i*_ is listed in Table 1.

**Table 1.** Confirmed cases of COVID-19 in the Republic of Korea and the corresponding deaths and fatality rates stratified by age groups as of 3/11/2020.

|  |  | Confirmed Cases n (%) |  | Deaths n (%) |  | Fatality Rate % |
| --- | --- | --- | --- | --- | --- | --- |
| Total |  | 8,413 | (100.0) | 84 | (100.0) | 1.0 |
| Age Groups | 80 and above | 286 | (3.4) | 31 | (36.9) | 10.84 |
|  | 70-79 | 542 | (6.44) | 29 | (34.52) | 5.35 |
|  | 60-69 | 1,059 | (12.59) | 16 | (19.05) | 1.51 |
|  | 50-59 | 1,615 | (19.2) | 6 | (7.14) | 0.37 |
|  | 40-49 | 1,171 | (13.92) | 1 | (1.19) | 0.09 |
|  | 30-39 | 873 | (10.38) | 1 | (1.19) | 0.11 |
|  | 20-29 | 2,342 | (27.84) | 0 | (0.0) | 0 |
|  | 10-19 | 438 | (5.21) | 0 | (0.0) | 0 |
|  | 0-9 | 87 | (1.03) | 0 | (0.0) | 0 |

If *V*_𝕋 𝔹_ *>* 1, then the population of 𝕋 has a higher risk of fatal outcomes due to a larger percentage of the older population. It results in a higher CFR compared to 𝔹. If *V*_𝕋 𝔹_ *<* 1, then 𝕋 has a younger population and it should result in a lower CFR compared to 𝔹.

Another correction factor is the estimated CFR of the benchmark country, *CFR*_𝔹_. For this manuscript we are using the latest known CFR as of 3/28/2020 from South Korea of *CFR*_𝔹_ = 0.01635.

The estimated cumulative case numbers *EC*(𝕋, *t*) at day *t* are directly computed from the estimated cumulative deaths that will occur 23 days later with respect to *V*_𝕋 𝔹_ and *CFR*_𝕋_.

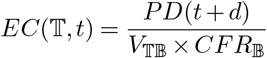

, for day shifting *d* = 23 representing the delay from infection to death.

## Results

Overlapping the reported exponential decrease from China, Italy, and Iran show a reproducible growth rate decline (Figure 2). We estimate the growth rate spline parameters *a* and *b* for *EDG*(*t*) with *a* = 0.4169 and *b* = 0.07073. This suggests that the growth rate after nationwide lockdown halves about every 10 days. By applying the proposed correction, the number of adjusted cases is significantly higher for most countries. Figure 3a shows the population age distribution of the US, Italy, and China compared to South Korea. Figure 3b shows the expected number of fatal outcomes for a 100% infection rate. The vulnerability factor for the US compared to South Korea is 1.07. This means that the population is equally vulnerable to fatal outcomes of COVID-19 infections. Italy, in contrast, has a vulnerability factor of 1.57. This is due to a higher fraction of the population being at a higher risk of death. This would indicate the expected CFR would be 57% higher in Italy compared to South Korea. China, with a younger population relative to South Korea, has a vulnerability factor of 0.63. The expected CFR in China should be lower than in South Korea based on the population risk. After applying the case adjustment, we observe a significant increase in the number of COVID-19 infections. The discrepancy in reported CFRs in combination with favorable population scores in the case of China and Iran suggest a over-proportional number of unreported COVID-19 infections. Figure 4 shows the projections of cumulative deaths for eight countries. The US has a projected number of COVID-19 fatalities of more than 80,000 before growth is reduced and new deaths are rare. The hospitalization per day will increase exponentially and reach its maximum in 12 to 14 days from the time of writing. Similar dynamics can be observed for other European countries. Table 2 shows the predicted number of cases and the Vulnerability Factor for seven countries. It suggests current infections in the US and Spain have exceeded 1.4 million.

**Table 2.** Reported and adjusted cases adjusted by South Korean CFR and Vulnerability Factor for 3/28/2020.

|  | Reported Cases | VF | Adjusted Cases |
| --- | --- | --- | --- |
| China | 82,198 | 0.69 | 325,452 |
| France | 45,170 | 1.38 | 676,839 |
| Italy | 101,739 | 1.57 | 965,113 |
| Spain | 87,956 | 1.36 | 1,421,505 |
| US | 161,807 | 1.07 | 1,417,635 |
| Germany | 66,885 | 1.48 | 222,068 |
| UK | 22,453 | 1.24 | 316,556 |

**Fig. 2.**
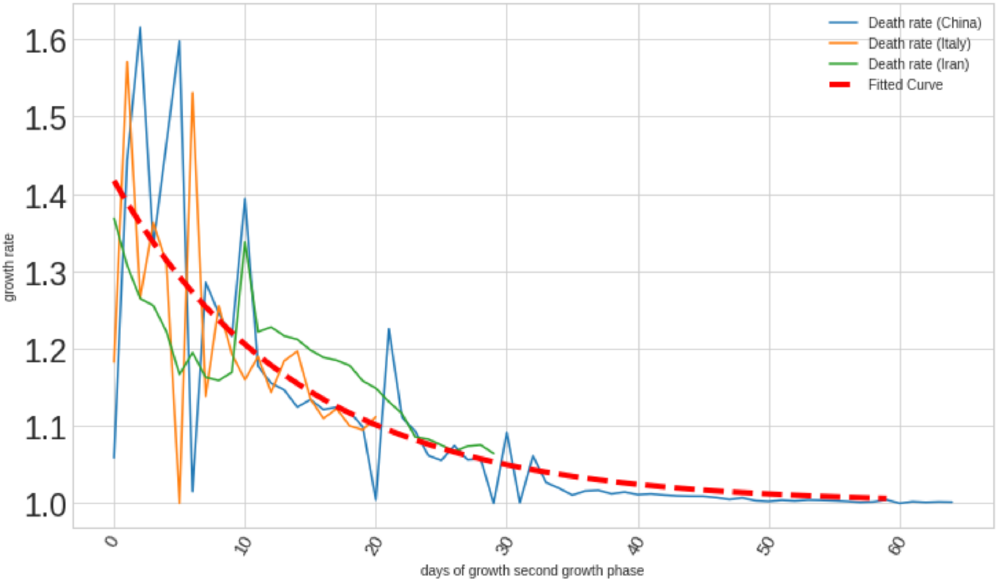
Matching exponentially decreasing growth rates fo countries undergoing growth rate modulating behavior change. The growth rate spline in red is the expected decrease of growth rate after lockdown measures applicable to other countries.

**Fig. 3.**
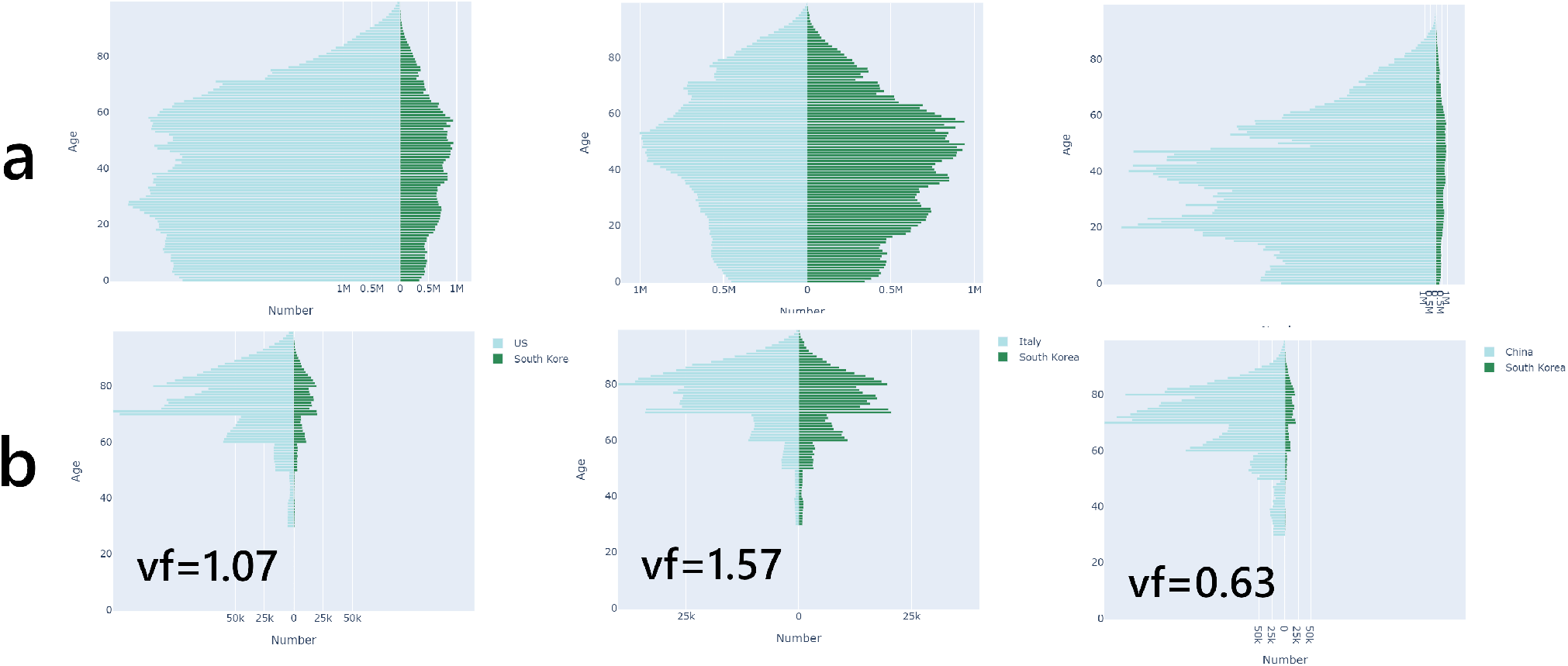
**a)** Population demographic comparison between US, Italy, and China compared to South Korea. **b)** Population demographic with fatal outcome of COVID-19 at 100% infection rate based on Table 1. Vulnerability Factor *V*_TB_ relative to South Korean population (vf).

**Fig. 4.**
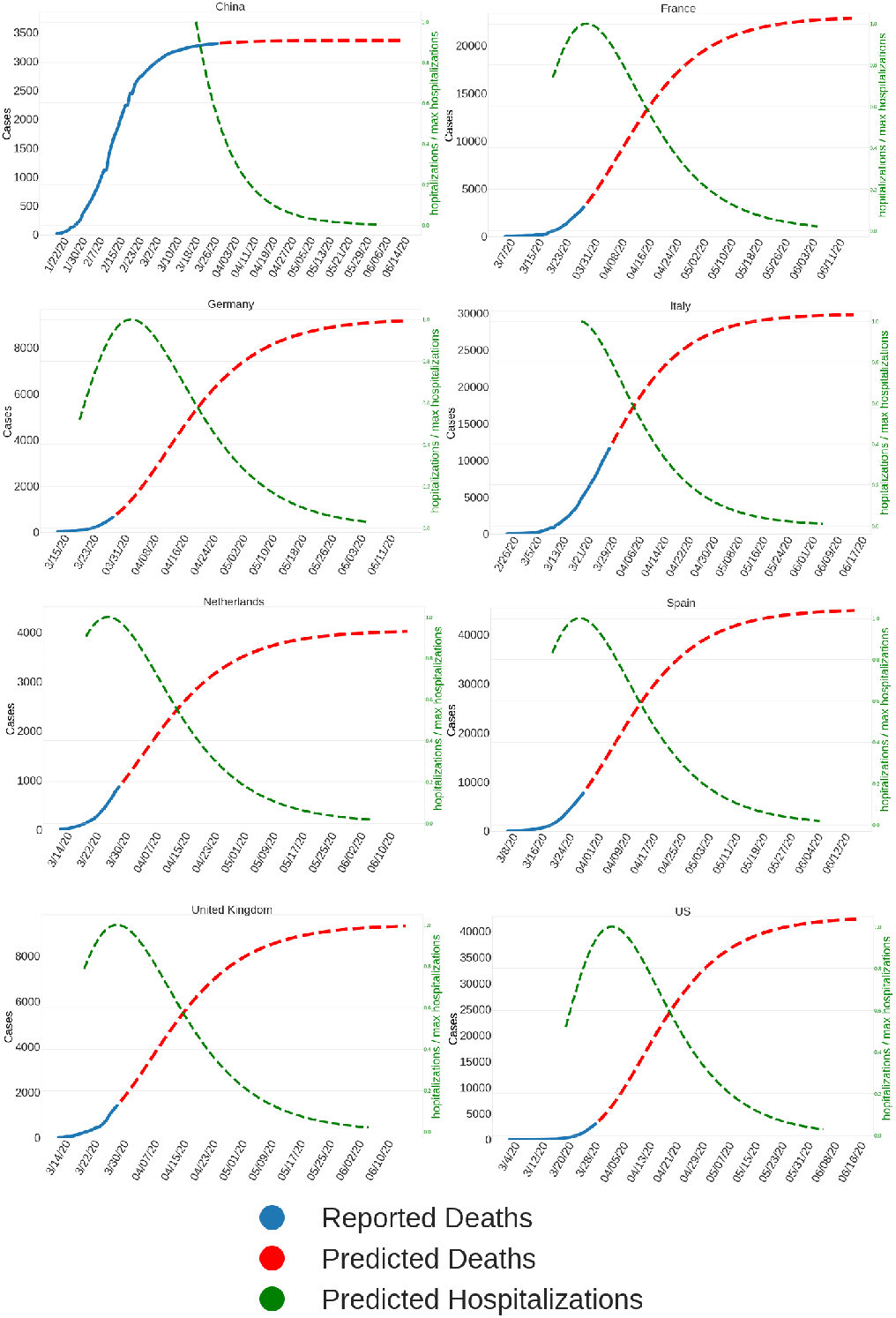
Predicted fatalities and expected hospitalizations for eight countries with major COVID-19 outbreaks. The left y-axis is the number of reported cumulative predicted fatalities. The right axis is the relative predicted daily hospitalizations normalized to the expected maximum of hospitalizations.

## Summary

The exponential growth curves indicate an extreme rise in fatalities world-wide. Data from China, Italy, and Iran suggest that extreme measures such as curfews and lockdown on national level can reduce the growth rate of COVID-19 over time. But even then the rate only decreases over a period of more than 30 days resulting in a significant number of new infections. It should not be assumed that the number of deaths drops quickly after such measures are taken. By extrapolating the cumulative deaths it is possible to project daily hospitalizations. The model proposed here does assume that only a fraction of the population is affected by the virus and growth limiting effects from a shrinking susceptibility pool are not the dominant factor. In cities like New York city we estimate that a substantial fraction of the population was infected or will be infected. The idea of flattening the curve (14) was not successful in New York City.

This study suggests that the current reporting of COVID-19 cases significantly underestimates the true scale of the pandemic. The lack of testing complicates the estimation of the true CFR and causes significant misinformation. This study has leveraged information derived from a well-tested sub-population (South Korea). With testing capacities of 20,000 tests daily, it has the largest and most accurate coverage compared with all other countries as of writing. The low false-negative rate in detecting COVID-19 infections leads to the lowest CFR compared with all other countries except Germany (1.49) with major case count. By applying the parameters estimated from this benchmark country, the proposed method can adjust global COVID-19 case numbers. This method is limited in its ability to predict the exact number of cases accurately. The requirement of predicting the exponential growth phase of the viral spread for a 23 day period will lead to high uncertainties, specifically to the onset of behavioral change. This method tries to apply a best case scenario in which lockdown measures are taking effect on the growth rates of cumulative deaths immediately. The method relies on the assumption that deaths by COVID-19 are detected and reported reliably. False-negative rates can have a distorting effect on the case adjustment. This is especially true if the benchmark country does not adequately report deaths from COVID-19. Additionally, the assumption of a globally similar CFRs is untested and should be applied with caution. The major finding in this study is a reproducible exponential decay of the growth rate. This exponential decrease is suggesting multiple doublings of viral infection after lockdown measures are taken. The model does not take population saturation into account such as SIR models. In the case of New York city, were a substantial proportion of the population might be infected, the model can over-estimate the growth rates. The predicted exponential decay function *EDG*(*t*) can be combined with SIR models to more accurately predict COVID-19 dynamics. This method suggests that due to the fast exponential growth of true case counts, most modern healthcare systems are not able to track the changes with sufficient and constant coverage. The model predicts a spike in hospitalizations at a rate that will add significant burden to healthcare ICU capacity in the next two weeks. It also highlights the importance of publicly accessible real-time data and the relevance of combining global healthcare efforts.

## Data Availability

All code and data is available on the Kaggle platform.

https://www.kaggle.com/lachmann12/correcting-under-reported-covid-19-case-numbers

https://github.com/lachmann12/covid19

## ACKNOWLEDGEMENTS

We want to thank Dr Avi Ma’ayan for feedback on the original manuscript and Alon Bar Tal for insightful discussion as well as the Kaggle community. Special thanks to the seamless accessibility of up-to-date COVID-19 case statistics published on GitHub by Johns Hopkins and the World Health Organization.

